# Prediction and Screening for Asymptomatic Carotid Artery Stenosis in Post-Radiotherapy Nasopharyngeal Carcinoma Patients

**DOI:** 10.1101/2025.08.19.25334024

**Authors:** Chuan-Yi Lin, Chun-Nan Chen, Jenq-Yuh Ko, Szu-Yuan Wu, Po-Hsiu Kuo

## Abstract

**Objective:** Head and neck radiotherapy (RT) is associated with an increased risk of carotid artery stenosis; yet standardized surveillance guidelines for nasopharyngeal carcinoma (NPC) survivors remain lacking. This study aimed to develop a risk prediction model to identify individuals at heightened risk.

**Methods:** We conducted this retrospective cohort study using claims data from the National Taiwan University Hospital-integrative Medical Database (NTUH-iMD). A Cox-based prediction model using stepwise variable selection was developed and model performance was evaluated using the area under receiver operating characteristic (ROC) curre (AUC) and integrated Brier score (IBS). To identify the optimal timing for initiating carotid ultrasound screening in post-RT NPC patients, a multivariable Generalized Linear Mixed Model (GLMM) was used to determine the post-treatment year most strongly associated with the development of moderate or greater internal carotid artery (ICA) stenosis.

**Results:** Patients in the high-risk group (risk score≥5) have significantly increased risk of moderate or greater ICA stenosis, which may begin to manifest as early as the fourth year after receiving radiation therapy. Additionally, we found that, compared to the average risk of the entire cohort, the risk of developing moderate-to-severe ICA stenosis began to increase significantly and persistently around the seventh year after radiotherapy. Moreover, the model achieved an AUC of 0.623 and an IBS of 0.084, indicating moderate discriminatory ability and good overall predictive accuracy.

**Conclusion:** According to our proposed risk prediction model, we recommend that carotid ultrasound screening begin in the seventh year following radiation therapy, while high-risk patients should start screening earlier, beginning in the fourth year.

## INTRODUCTION

### Nasopharyngeal carcinoma and radiotherapy

Nasopharyngeal carcinoma (NPC) is a relatively rare malignancy worldwide; however, it exhibits a higher prevalence in certain endemic regions, including Taiwan, Southern China, Hong Kong, and Singapore^1,2^. The annual incidence of NPC is about 6.17 per 100,000 in Taiwan^3^. NPC is a disease with highly sensitive ton radiation. Therefore, radiotherapy (RT) alone and concurrent chemo-radiotherapy (CCRT) are regarded as the most efficient treatment for early and advanced stages of disease^4^. In the modern era of intensity-modulated radiation therapy (IMRT) and effective systemic chemotherapy, NPC patients had favorable overall survival. The 5-year overall survival is reported to be 65.2% and 10-year overall survival is about 60% in Taiwan^5,6^. However, after irradiation, the late complications, such as cranial nerve palsy, endocrine dysfunction, dysphagia and carotid artery disease^7^ significantly affect patients’ quality of life. With larger number of long-term survivors, the prediction and prevention of late RT-related complications has become important.

### Head and neck irradiation increase risk of carotid artery stenosis and ischemic stroke

Ischemic stroke is a known late complication of neck irradiation^3,8^. RT may trigger pro-inflammatory responses that increase endothelial permeability, promote inflammatory cell infiltration, and accelerate atherosclerosis^9,10^. Additional mechanisms of radiation-induced vasculopathy include vasa vasorum injury and adventitial fibrosis, which impair vessel wall perfusion and increase vascular stiffness, ultimately contributing to progressive carotid artery stenosis (CAS)^11,12^. CAS is defined as atherosclerotic plaque development in the internal carotid artery (ICA). The plaque can cause narrowing of the carotid artery and blocking blood flow which may lead to an increased risk of cerebrovascular events. CAS can be classified as mild (<50%), moderate (≥50%) or severe (≥70%) according to the percentage of blockage and the severity of the stenosis. According to past studies, NPC patients who had received RT have higher prevalence of carotid plaque and greater degree of CAS than patients who have not received RT^13,14^. In addition, CAS is a well-established and clinically significant risk factor for ischemic stroke, accounting for up to 20% of all ischemic strokes or transient ischemic attacks^15^. Lee et al. reported a 1.66-fold adjusted increase in stroke risk among younger (<55 years old) survivors of NPC who underwent RT or CCRT^5^. Furthermore, the onset of stroke among NPC survivors occurs approximately 10 years earlier than in the general population^16^, highlighting the accelerated cerebrovascular aging and elevated long-term vascular risk in this patient group.

### Screening for carotid artery stenosis in high-risk populations is cost-effectiveness

The prevalence of CAS increases with age and in the presence of risk factors such as hypertension, dyslipidemia, diabetes mellitus, smoking, obesity, peripheral arterial occlusive disease, coronary artery disease (CAD), and other cardiac conditions^17^. Patients diagnosed with significant stenosis should be considered for appropriate medical management and/or carotid intervention.^18^ In addition, screening asymptomatic populations with a 20% prevalence of CAS of 60% or greater has been shown to result in a net stroke reduction, preventing approximately 7.9 strokes per 1,000 individuals screened^19^. This evidence supports the potential clinical benefit of targeted screening programs in high-risk groups to identify significant CAS and implement timely preventive interventions. Carotid Doppler ultrasonography (US) is currently the most appropriate method for assessment of the ICA because of its radiation free, availability and less invasive property^20^. Therefore, screening of CAS by using carotid Doppler US appears to play an important role in preventing ischemic stroke in post-RT NPC patients. This study aimed to identify risk factors for CAS in patients with NPC, develop a risk prediction model for moderate or greater ICA stenosis, and determine the post-radiotherapy period most strongly associated with stenosis progression using carotid Doppler US.

## METHODS

### Data source and study population

We conducted this retrospective cohort study using claims data from the National Taiwan University Hospital-integrative Medical Database (NTUH-iMD). The NTUH-iMD systematically records structured data (such as laboratory results and diagnoses), semi-structured data (including medication orders), and unstructured data (such as clinical notes) in standardized formats, thereby ensuring consistency and reliability for research and analysis. Diagnoses were documented using International Classification of Diseases (ICD) Clinical Modification codes. Specifically, the Ninth Revision (ICD-9-CM) was utilized prior to 2016, after which the Tenth Revision (ICD-10-CM) was implemented starting in 2016.

Enrolled patients in this study based on following criteria: (1) age 18 years or older (2) diagnosed with NPC (ICD-9 149; ICD-10 C.11) between 2008 and 2018 and the diagnosis of NPC was confirmed with NTUH Cancer Registry database or pathological report (3) received standard treatment according to clinical stage based on the 8th edition of the AJCC (4) had undergone more than two carotid Doppler US examinations after being diagnosed with NPC between 2008 and 2023. The exclusion criteria were as follows: (1) previous stroke history, including infarction/hemorrhage/lacunar infarction (2) previous acute myocardial infraction history (3) history of radiotherapy (4) previous history of malignancy or diagnosis of second malignancy during follow-up period (5) NPC recurrence observed during follow-up period (6) declined to undergo treatment or lost follow-up after diagnosed of NPC. This study was approved by the Research Ethics Committee of NTUH and complied with the Declaration of Helsinki (approval number: 202403116RINE).

### Primary outcome and follow-up time

The primary outcome was defined as the development of moderate or greater (≥50%) ICA stenosis during the follow-up period, which extended until December 31, 2023.

### Risk factors of carotid artery stenosis

The index date was defined as the patient diagnosed NPC. Several risk factors that can result in CAS were adjusted to interpretate for potential confounding effects. The adjusted covariates included age, sex, body mass index (BMI), cigarette smoking, and coexisting comorbidities associated with the primary outcome, such as diabetes, hypertension, dyslipidemia, CAD, and chronic kidney disease. The ICD-9-CM code and ICD-10-CM code were used to document and categorize these comorbidities, starting two years forward from the index date. In addition, we examined prescriptions of anti-thrombotic drugs such as aspirin, clopidogrel, rivaroxaban, apixaban, edoxaban, betrixaban, and warfarin during the same period.

### Statistical analysis

A multivariable Cox proportional hazards (PH) model was constructed to assess associations between risk factors and the development of moderate or greater ICA stenosis, using an Akaike Information Criterion (AIC)-based backward stepwise selection procedure^21^. The final model was applied to estimate individual risk, and a risk score was derived from the model coefficients to facilitate risk prediction. Points assigned to each risk factor category were summed to calculate the total risk score for CAS progression. To develop the risk prediction model, 70% of the total cases were allocated to the training dataset, and the remaining 30% were designated as the validation dataset for cross-validation. Additionally, the full sample was used for bootstrapping to perform internal validation of the model to test the overfitting problem^22^. Receiver operating characteristic (ROC) curves were plotted, and area under the curves (AUC) were calculated as C statistic to assess the discrimination of the prediction model^23^. In addition, we calculated the integrated Brier score (IBS) to inspect the overall accuracy over time^24,25^. The IBS is an extension of the Brier Score designed to assess the accuracy of probabilistic predictions over time in survival analysis. Lower IBS values indicate higher overall prediction accuracy. An IBS of 0.2 or lower is generally considered acceptable for binary outcomes, with values between 0.1 and 0.2 reflecting good performance, and values between 0 and 0.1 indicating excellent performance. Furthermore, to evaluate the model’s ability to discriminate among different risk levels, a predictiveness curve was generated. The predictiveness curve was proposed by Pepe et al.^26^ and Huang et al.^27^ to describe the predictive capacity of a risk prediction model or marker. It displays the population distribution of risk via the risk quantiles and assesses the usefulness of a risk model when applied to the population. A well-performing model should successfully identify a high-risk subgroup suitable for targeted intervention. The flowchart outlining the methodology of this study is shown in Figure 1.

**Figure 1.**
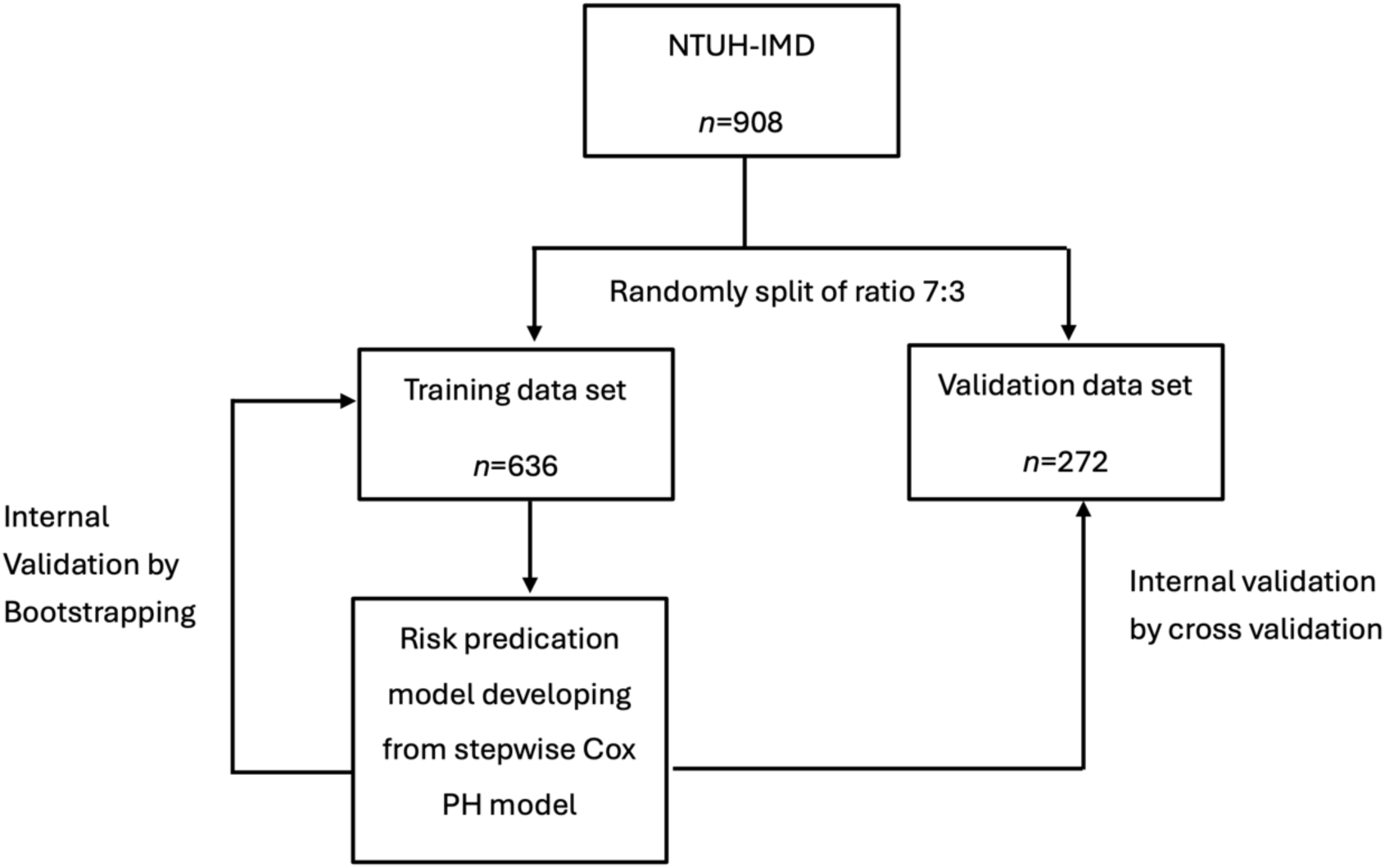
The Comprehensive Research Framework of This Study.

On the other hand, a multivariable generalized linear mixed model (GLMM) was applied to examine factors associated with the development of moderate or greater (≥50%) ICA stenosis following RT. The binary outcome was defined by the presence or absence of significant stenosis, as determined through serial carotid Doppler US examinations during follow-up. Fixed effects included age, sex, BMI, history of coronary artery disease, and time since RT (in years), with a patient-specific random intercept to account for within-subject correlation due to repeated measures. The model employed a logit link function and was specified as follows: logit(μ*ij*) = β0+β1 ⋅Age*ij*+β2⋅Gender*i*+β3⋅BMI*ij*+β4⋅CAD*i*+β5⋅Time*ij*+b*i* where μ*ij* the predicted probability of moderate ICA stenosis for patient *i* at time point *j*, and b*i*∼*N*(0, σ2) is the patient-specific random effect. Time since RT was modeled to characterize temporal risk trajectories and to identify periods of elevated risk, thereby informing the optimal timing for carotid ultrasound surveillance in post-RT NPC patients. All analyses in this study were realized using R version 4.2.1 (R Project for Statistical Computing) and SAS (version 9.4; SAS Institute, Inc., Cary, NC, USA)

## RESULTS

### Baseline characteristics of participants

A total of 908 cases met our inclusion criteria and were included in this study. The selection process of participants was shown in Figure. 1. Overall, the mean age of the participants was 48.51 ± 10.73. The mean follow-up was 7.40 ± 3.04 years and the mean time to outcome (≥50% ICA stenosis) was 8.00 ± 2.99 years. The baseline characteristics of enrolled patients was presented as Table 1.

**Table 1.** Characteristics of Nasopharyngeal Cancer Patients Enrolled in This Study.

|  | All<br>N=908 |  | <50% stenosis of ICA<br>N= 776 (85.5%) |  | >= 50% stenosis of ICA<br>N= 132 (14.5%) |  | P-value |
| --- | --- | --- | --- | --- | --- | --- | --- |
|  | N | % | N | % | N | % |  |
| **Age (mean ± SD)** | 48.51 ± 10.73 | | 48.37 ± 10.79 | | 51.02 ± 9.98 | | 0.0566 |
| **Age median, (IQR), years** | 48.19 (40.73,55.68) |  | 47.77 (40.38, 55.46) |  | 49.92 (43.85, 57.25) |  | 0.0311 |
| **Age group, years** |  |  |  |  |  |  | 0.0637 |
| <50 | 521 | 57.38% | 455 | 58.63% | 66 | 50.00% |  |
| >=50 | 387 | 42.62% | 321 | 41.37% | 66 | 50.00% |  |
| **Sex** |  |  |  |  |  |  | <.0001 |
| Female | 257 | 28.30% | 239 | 30.80% | 18 | 13.64% |  |
| Male | 651 | 71.70% | 537 | 69.20% | 114 | 86.36% |  |
| **BMI** |  |  |  |  |  |  | 0.0637 |
| Normal | 397 | 43.72% | 349 | 44.97% | 48 | 36.36% |  |
| Overweight | 424 | 46.70% | 359 | 46.26% | 65 | 49.24% |  |
| Obesity | 87 | 9.58% | 68 | 8.76% | 19 | 14.39% |  |
| **Comorbidities** |  |  |  |  |  |  |  |
| **Diabetes** | 866 | 95.37% | 744 | 95.88% | 122 | 92.42% | 0.0645 |
| No | 42 | 4.63% | 32 | 4.12% | 10 | 7.58% |  |
| Yes |  |  |  |  |  |  |  |
| **Hypertension** |  |  |  |  |  |  | 0.1629 |
| No | 827 | 91.08% | 711 | 91.62% | 116 | 87.88% |  |
| Yes | 81 | 8.92% | 65 | 8.38% | 16 | 12.12% |  |
| **Dyslipidemia** |  |  |  |  |  |  | 0.5059 |
| No | 851 | 93.72% | 729 | 93.94% | 122 | 92.42% |  |
| Yes | 57 | 6.28% | 47 | 6.06% | 10 | 7.58% |  |
| **Coronary artery disease** |  |  |  |  |  |  | <.0001 |
| No | 881 | 97.03% | 760 | 97.94% | 121 | 91.67% |  |
| Yes | 27 | 2.97% | 16 | 2.06% | 11 | 8.33% |  |
| **Chronic kidney disease** |  |  |  |  |  |  | 0.3551 |
| No | 903 | 99.45% | 771 | 99.36% | 132 | 100% |  |
| Yes | 5 | 0.55% | 5 | 0.64% | 0 | 0% |  |
| **Anti-thrombotic drugs** |  |  |  |  |  |  | 0.6918 |
| No | 853 | 93.94% | 730 | 94.07% | 123 | 93.18% |  |
| Yes | 55 | 6.06% | 46 | 5.93% | 9 | 6.82% |  |
| **Smoking** |  |  |  |  |  |  | 0.5713 |
| Never | 638 | 70.26% | 548 | 70.62% | 90 | 68.18% |  |
| Current/Former | 270 | 29.74% | 228 | 29.38% | 42 | 31.82% |  |
| **Follow-up year,**<br>**(mean ± SD)** | 7.40 ± 3.04 |  | 7.30 ± 3.03 |  | 8.00 ± 2.99 |  | 0.0147 |

**ABBREVIATIONS:** ICA, internal carotid artery; SD, standard deviation; IOR, interquartile range; BMI, body mass index

A total of 132 individuals (14.5%) developed moderate or greater ICA stenosis during the follow-up period. Compared to those without significant ICA stenosis, affected individuals were notably older (51.02 ± 9.98 vs. 48.37 ± 10.79 years), more likely to be male (86.36% vs. 69.20%), and had a higher prevalence of obesity (14.39% vs. 8.76%). No significant differences were observed between the two groups in terms of cigarette smoking or use of antithrombotic medications. Additionally, comorbidities such as diabetes, hypertension, dyslipidemia, and chronic kidney disease did not differ significantly (P > 0.05). However, a higher proportion of individuals in the moderate or greater ICA stenosis group had a history of CAD (82.0% vs. 73.2%) (Table 1).

### Risk prediction model for moderate or greater ICA stenosis in post-RT NPC patients

Based on multivariate Cox PH models conducted within the training dataset (n=636), Table 2 showed risk factors for moderate or greater ICA stenosis. We established a risk predication model based on stepwise Cox PH model. Six variables, including age, gender, BMI (overweight/obesity), CAD and smoking habits were selected in our prediction model. Predicted risks of moderate or greater ICA stenosis were derived from the cumulative scores of these variables (Table 3). Participants were categorized into three groups according to their scores: low risk (<3 points), moderate risk (3–4 points), and high risk (≥5 points). The Kaplan–Meier curves showed favorable discriminatory capacity of the model, with log-rank p < 0.001 (Figure 2). The high-risk group showed the highest risk of developing moderate or greater ICA stenosis and the risk increases significantly starting in the fourth year after RT. When significant ICA stenosis occurs, the high-risk groups are the earliest, followed by the moderate-risk group and the low-risk group.

**Figure 2.**
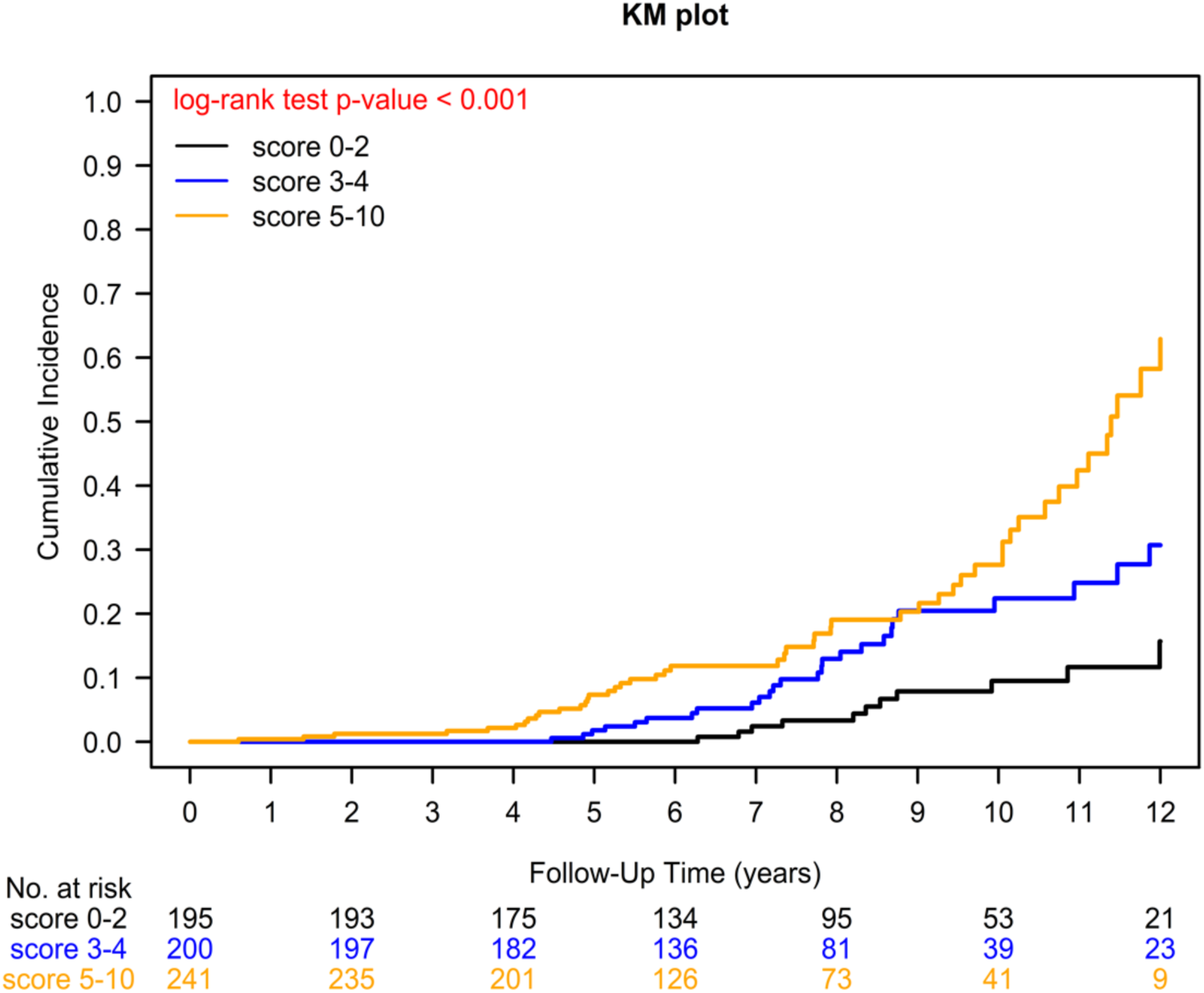
Kaplan-Meier Analysis of Cumulative Incidence of Moderate or Greater Internal Carotid Artery Stenosis in Post-Radiotherapy Nasopharyngeal Carcinoma Patients.

**Table 2.**
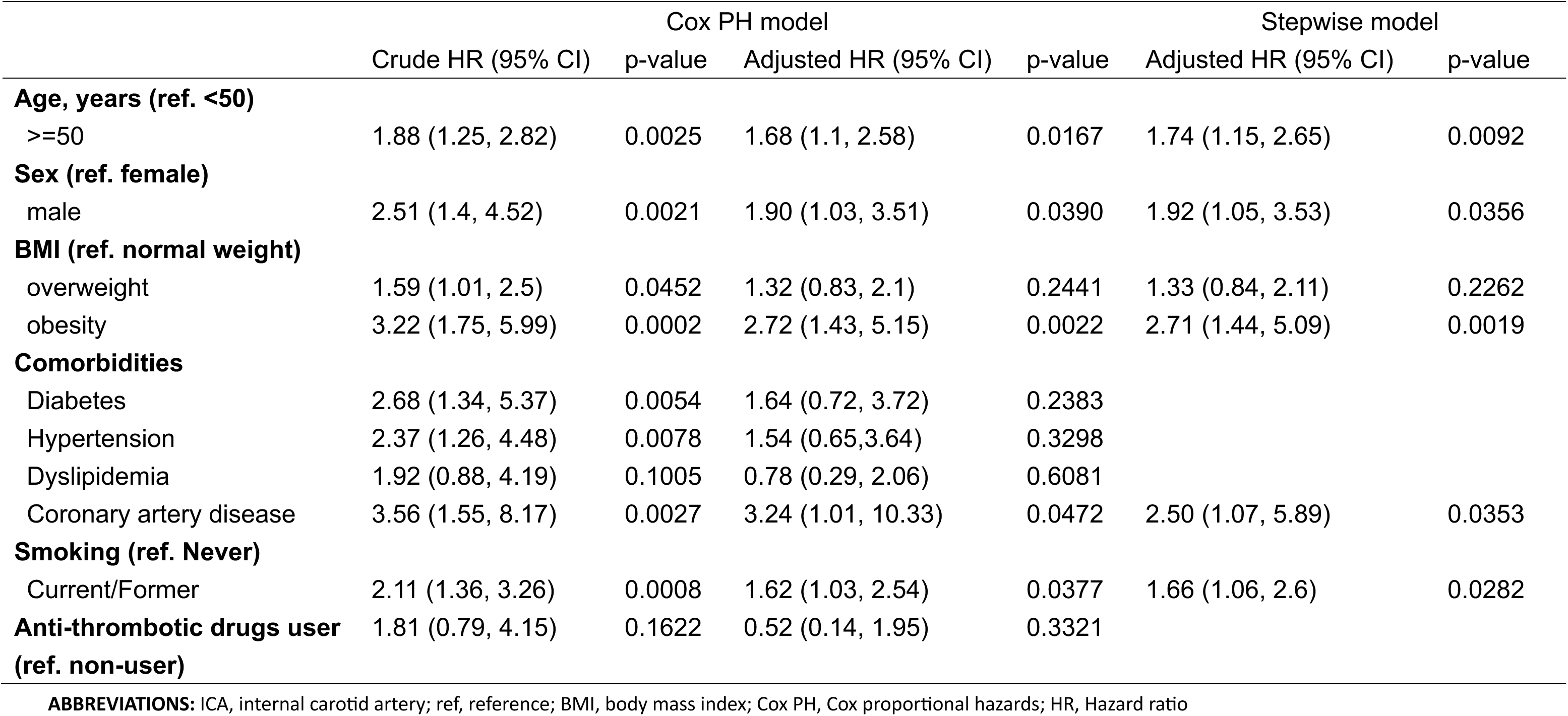
Risk Factors for Moderate or Greater ICA Stenosis in Post Radiotherapy Nasopharyngeal Cancer Patients (N=636)

|  | Cox PH model |  |  |  | Stepwise model |  |
| --- | --- | --- | --- | --- | --- | --- |
|  | Crude HR (95% CI) | p-value | Adjusted HR (95% CI) | p-value | Adjusted HR (95% CI) | p-value |
| **Age, years (ref. <50)** |  |  |  |  |  |  |
| >=50 | 1.88 (1.25, 2.82) | 0.0025 | 1.68 (1.1, 2.58) | 0.0167 | 1.74 (1.15, 2.65) | 0.0092 |
| **Sex (ref. female)** |  |  |  |  |  |  |
| male | 2.51 (1.4, 4.52) | 0.0021 | 1.90 (1.03, 3.51) | 0.0390 | 1.92 (1.05, 3.53) | 0.0356 |
| **BMI (ref. normal weight)** |  |  |  |  |  |  |
| overweight | 1.59 (1.01, 2.5) | 0.0452 | 1.32 (0.83, 2.1) | 0.2441 | 1.33 (0.84, 2.11) | 0.2262 |
| obesity | 3.22 (1.75, 5.99) | 0.0002 | 2.72 (1.43, 5.15) | 0.0022 | 2.71 (1.44, 5.09) | 0.0019 |
| **Comorbidities** |  |  |  |  |  |  |
| Diabetes | 2.68 (1.34, 5.37) | 0.0054 | 1.64 (0.72, 3.72) | 0.2383 |  |  |
| Hypertension | 2.37 (1.26, 4.48) | 0.0078 | 1.54 (0.65, 3.64) | 0.3298 |  |  |
| Dyslipidemia | 1.92 (0.88, 4.19) | 0.1005 | 0.78 (0.29, 2.06) | 0.6081 |  |  |
| Coronary artery disease | 3.56 (1.55, 8.17) | 0.0027 | 3.24 (1.01, 10.33) | 0.0472 | 2.50 (1.07, 5.89) | 0.0353 |
| **Smoking (ref. Never)** |  |  |  |  |  |  |
| Current/Former | 2.11 (1.36, 3.26) | 0.0008 | 1.62 (1.03, 2.54) | 0.0377 | 1.66 (1.06, 2.6) | 0.0282 |
| **Anti-thrombotic drugs user (ref. non-user)** | 1.81 (0.79, 4.15) | 0.1622 | 0.52 (0.14, 1.95) | 0.3321 |  |  |

**ABBREVIATIONS:** ICA, internal carotid artery; ref, reference; BMI, body mass index; Cox PH, Cox proportional hazards; HR, Hazard ratio

**Table 3.**
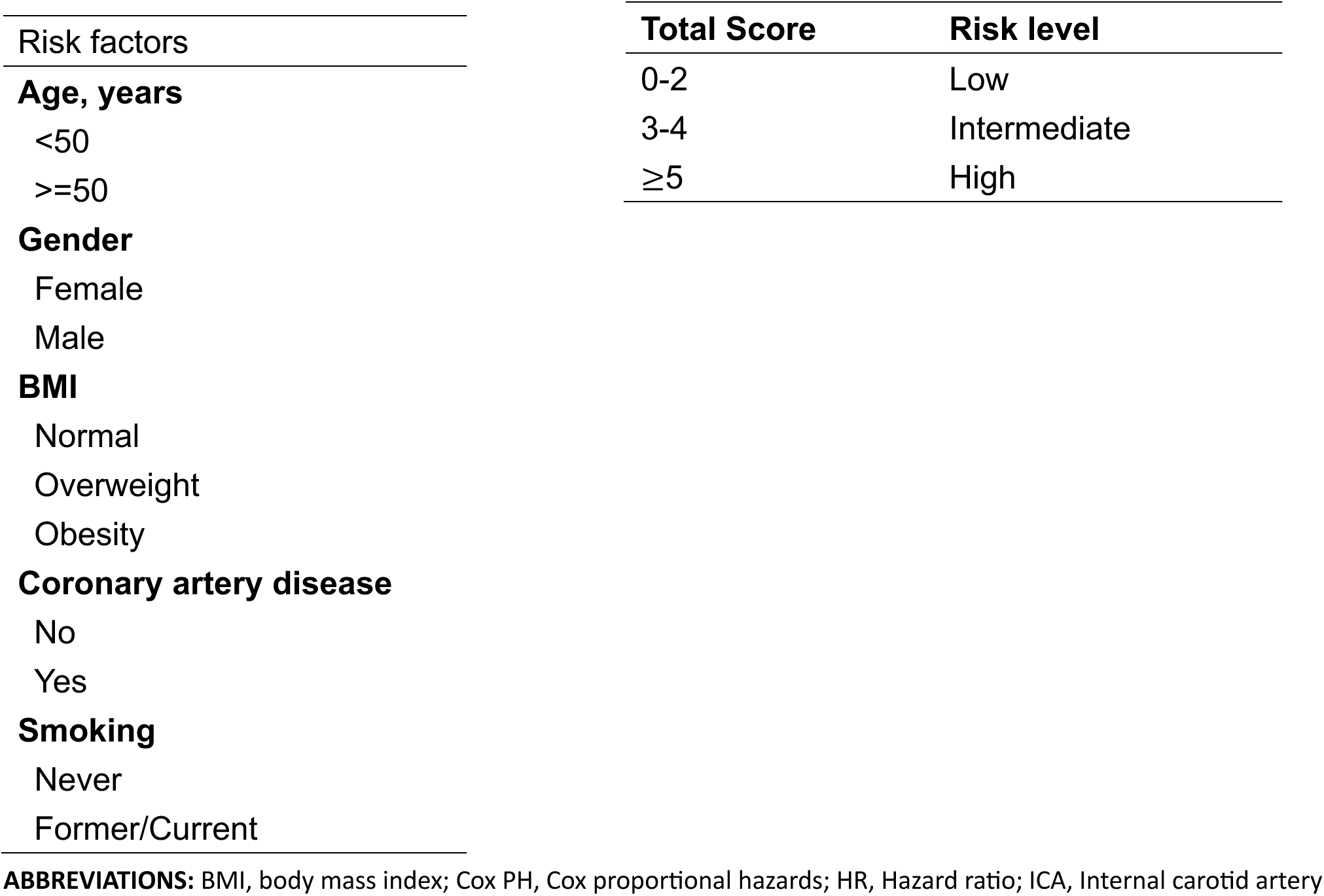
Risk Scores and Risk Prediction Model for Moderate or greater ICA Stenosis in Post-Radiotherapy Nasopharyngeal Cancer Patients from Stepwise Cox PH model.

### Performance of this risk prediction model

The area under ROC curve (AUC) of the model for predicting moderate or greater ICA stenosis was 0.623 (Figure 3), indicating a moderate ability to discriminate between patients with and without significant ICA stenosis. Internal validation was performed using bootstrap resampling of all participants (n = 908), which yielded an AUC of 0.615. In addition, cross-validation was conducted using 30% of the enrolled cases (n = 272), and a moderate level of discrimination was observed (AUC = 0.595). This result suggest that the model demonstrates consistent, though moderate, discriminative ability across different validation approaches and no correction for over-optimism in the beta coefficients was necessary. Furthermore, the Delong test was done, and there was no significant difference in the predictability of this pretested model between the training cohort and validation cohort (Figure 3). To further assess the model’s discriminative ability across different risk strata, a predictiveness curve was constructed. As shown in the Figure 4, the predicted risk increases gradually across the lower and middle percentiles but rises sharply in the upper percentiles— particularly beyond the 90th percentile. This right-skewed distribution indicates that the model effectively concentrates predicted risk among a small subset of individuals. Such stratification suggests that the model has strong discriminatory capacity and may be useful for identifying a high-risk subgroup that could benefit from targeted preventive or therapeutic interventions.

**Figure 3.**
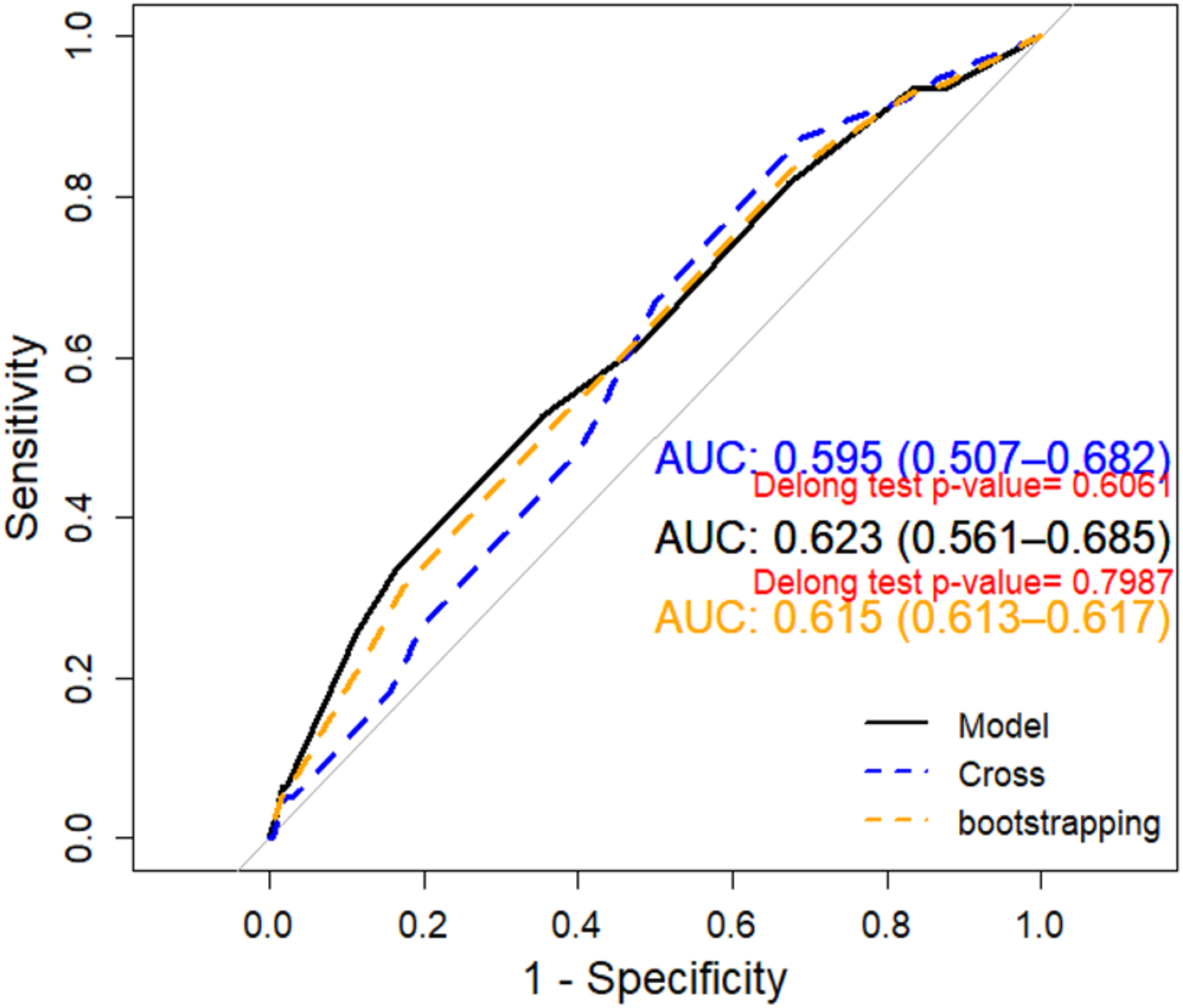
The ROC Curve of Risk Prediction Model with Bootstrapping Validation and Cross Validation.

**Figure 4.**
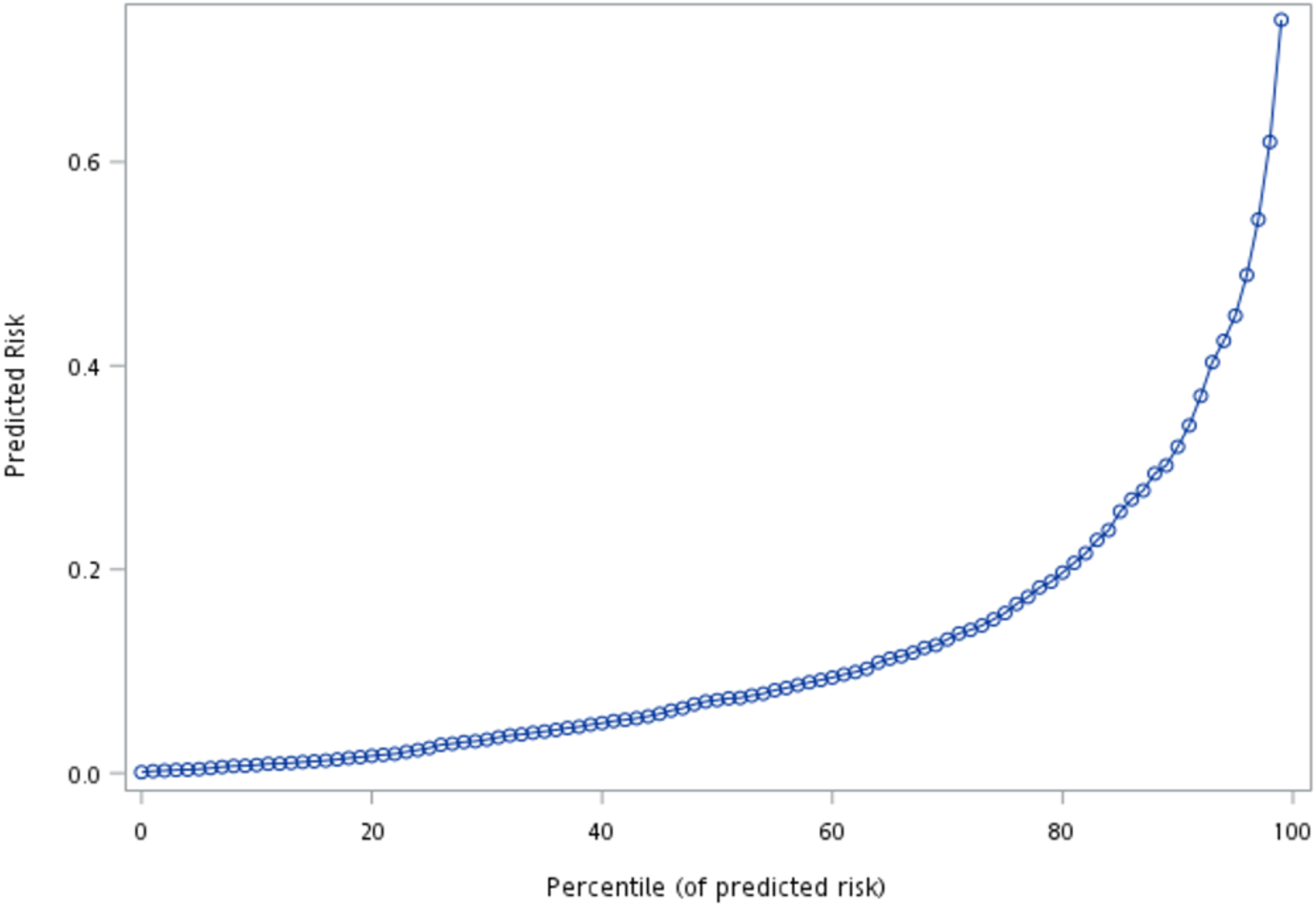
The Predictiveness Curve of Risk Prediction Model. The curve shows the predicted risk of moderate or greater internal carotid artery (ICA) stenosis across percentiles of the predicted risk distribution. A steep increase in predicted risk is observed in the upper percentiles, suggesting strong discrimination and the model’s ability to identify a high-risk subgroup.

In addition, the IBS was calculated to assess the model’s average predictive accuracy over the entire follow-up period. The resulting IBS of 0.084 indicates good overall performance, reflecting adequate calibration and time-dependent predictive accuracy for moderate-to-severe ICA stenosis. This value falls within the acceptable range for survival models, suggesting the model effectively captures the temporal dynamics of stenosis progression following radiotherapy.

### Time to onset of significant ICA stenosis post-radiotherapy

The GLMM identified several significant predictors for moderate or greater ICA stenosis following RT. The results are summarized in Table 4. A positive correlation was observed between the duration after radiation therapy and the severity of ICA stenosis. The risk of moderate or greater ICA stenosis increased with time elapsed since radiation. When compared to patients within 7 years post-radiation, those at 8 to 11 years post-radiation had a significantly higher risk (Odds ratio, OR = 3.98; 95% CI, 2.94–5.38; P < .0001), while patients beyond 12 years post-radiation exhibited an even greater risk (OR = 8.81; 95% CI, 5.86–13.26; P < .0001). In addition, age ≥50 years was associated with an almost two-fold increase in risk (OR=1.96; 95% CI, 1.45–2.64; P < .0001), consistent with age-related vascular changes. Male sex was a significant predictor (OR= 2.17; 95% CI, 1.47–3.20; P < .0001), suggesting sex-based vulnerability to radiation-induced vascular injury. Obesity (BMI ≥30) significantly increased the odds of ICA stenosis compared with normal weight (OR= 2.05; 95% CI, 1.28–3.28; P = 0.002), highlighting the role of central obesity as a vascular risk factor. Interestingly, overweight status (BMI 25– 29) was not associated with increased risk (OR= 1.07; P = 0.669). Furthermore, a history of CAD was also strongly associated with ICA stenosis (OR= 2.38; 95% CI, 1.57–3.61; P < .001), reinforcing the link between systemic atherosclerosis and radiation-induced vascular complications. Smoking history was another independent predictor (OR= 1.60; 95% CI, 1.18–2.16; P = 0.002), underscoring the additive vascular damage from tobacco exposure.

**Table 4.**
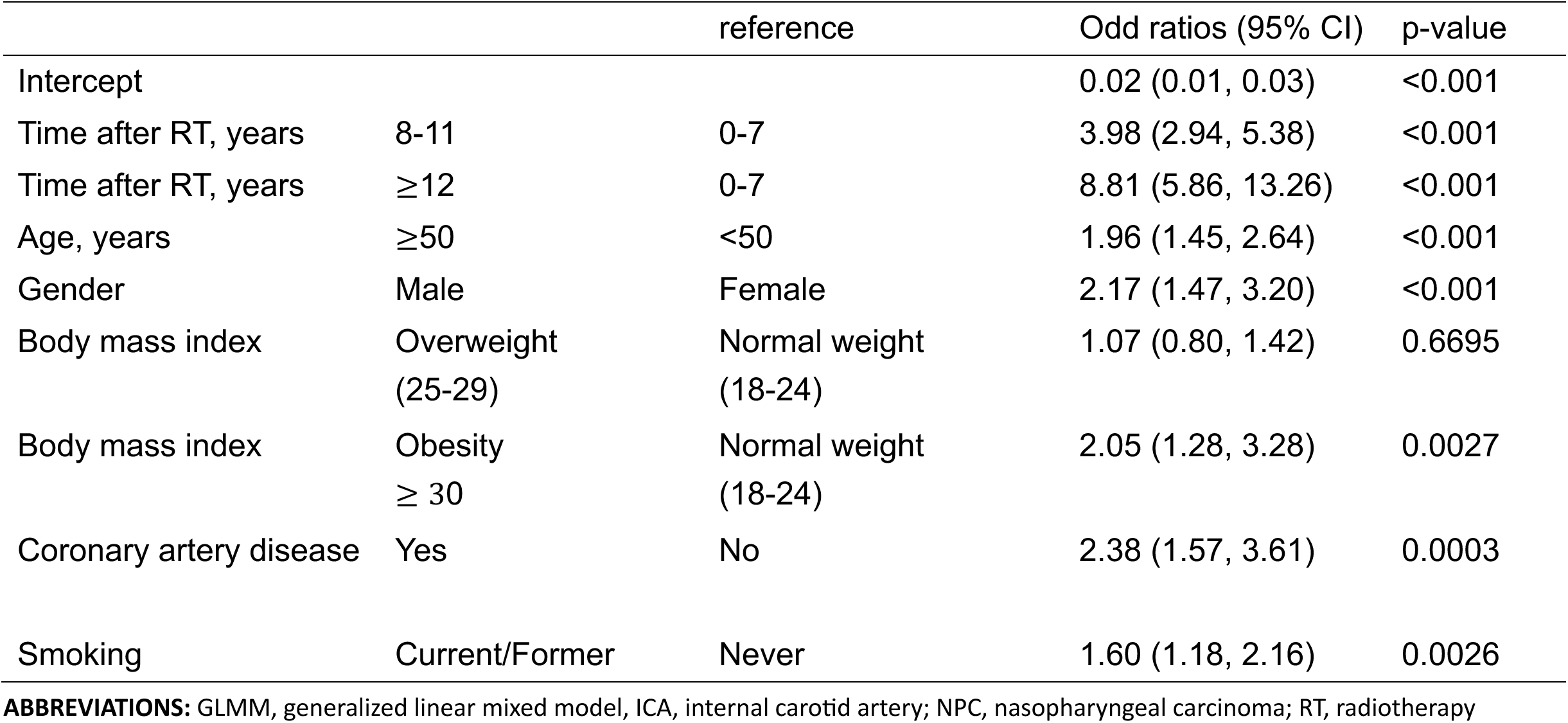
Multivariable GLMM Analysis of Factors Associated with Moderate or Greater ICA Stenosis in post-RT NPC patients.

As shown in the Figure 5, we found that, compared to the average risk of the entire cohort, the risk of developing moderate-to-severe ICA stenosis began to increase significantly and persistently around the seventh year after radiation therapy. In other words, there is a time-dependent increase in the odds of developing internal carotid artery stenosis in patients with NPC after receiving RT. These findings emphasize that long-term surveillance and monitoring guidelines should be specifically established for patients who are older than 50 years, male, obese, have a history of smoking, or have pre-existing CAD.

**Figure 5.**
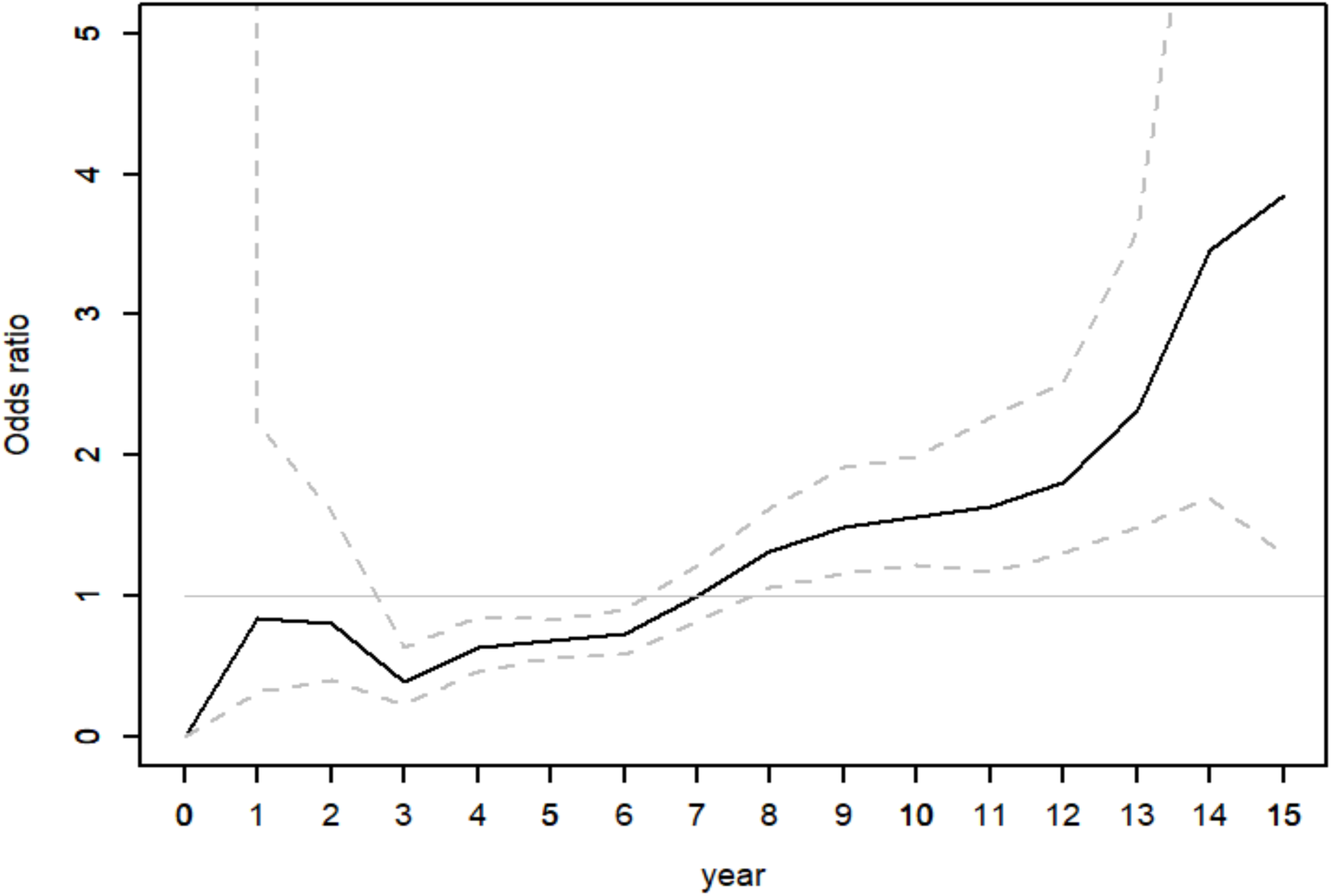
Time-Dependent Increase in Odds of Internal Carotid Artery Stenosis Following Radiotherapy in Patients with Nasopharyngeal Carcinoma. The solid line represents the estimated annual odds ratio (OR) for internal carotid artery (ICA) stenosis following radiotherapy in patients with nasopharyngeal carcinoma (NPC), while the dashed lines indicate the 95% confidence interval. The horizontal reference line at OR = 1 reflects the average post-radiotherapy ICA stenosis risk in the entire NPC cohort. A marked increase in risk is observed beginning at year 7, suggesting a long-term vascular effect of radiotherapy.

## DISCUSSION

### The main findings of this study

Based on our proposed risk prediction model, patients in the high-risk group (risk score≥5) have significantly increased risk of moderate or greater ICA stenosis, which may begin to manifest as early as the fourth year after receiving RT. The enrolled participants had a mean age of 48.5 years and were followed for an average of 7.4 years. During the follow-up period, 14.5% of participants developed moderate or greater ICA stenosis. Compared to previous studies^28,29^, the prevalence of moderate or greater ICA stenosis in our study population was markedly higher than that reported in the general population of the same age group. This further confirms that receiving radiation therapy to the head and neck substantially increases the risk of CAS. Moreover, in the analysis using a GLMM, we found that, compared to the average risk of the entire cohort, the risk of developing moderate or greater ICA stenosis began to increase significantly and persistently around the seventh year after radiation therapy (Figure 5). This finding suggests that routine carotid ultrasound screening and follow-up should be initiated from the seventh year onward for all NPC patients who have undergone RT.

### The pathophysiology of radiotherapy induced carotid atherosclerosis

Radiotherapy can induce tissue necrosis and inflammation, subsequently leading to endothelial injury, cellular proliferation, and fibrosis^30,31^. These processes, along with accelerated atherosclerosis, are considered key mechanisms underlying radiation-induced vascular injury. Notably, these mechanisms closely align with those proposed in Virchow’s theory^32^. Certainly, ionizing radiation can cause direct and immediate injury to endothelial cells, disrupting their structural integrity and compromising vascular permeability. This endothelial damage triggers a cascade of inflammatory responses, which contribute to vascular dysfunction and the development of radiation-induced tissue injury. The oxidation of LDL molecules is promoted by the presence of free radicals, which initiate oxidative modifications that transform LDL into a more atherogenic form. This oxidized LDL plays a crucial role in the pathogenesis of atherosclerosis by triggering inflammatory responses and foam cell formation within the arterial wall. The formation of foam cells—which persist within the intima for extended periods—is a hallmark of RT-induced vascular damage.

### Identifying risk factors for significant ICA stenosis in post-RT NPC patients using our predictive model

Our study demonstrated that male patients were more prone to developing moderate or greater ICA stenosis compared to female patients, adjusted HR= 1.90 (95% CI: 1.03-3.51). Moreover, individuals aged over 50 showed a significantly higher incidence of significant ICA stenosis, adjusted HR=1.68 (95% CI: 1.1-2.58). These findings consistent with previous studies conducted in the general population^33,34^. Interestingly, we observed that NPC patients who had undergone RT developed moderate or greater ICA stenosis at a notably younger average age than that reported in the general population. This suggests that radiation exposure may accelerate vascular aging or atherosclerotic changes, underscoring the need for earlier screening strategies in this high-risk group. In the multivariate Cox regression analysis, the comorbidities such as hypertension, diabetes, and hyperlipidemia did not show a significant impact to outcome. This may be attributed to the relatively young average age of the study population, as the duration of these chronic diseases may not have been long enough to contribute significantly to the development of carotid artery stenosis. Alternatively, factors such as CAD and obesity may be more strongly associated with CAS than the aforementioned chronic diseases. The association between carotid atherosclerosis and CAD has been well established^35–37^. CAD reflects pre-existing atherosclerotic changes in the vessels supplying the heart and atherosclerosis is considered to be a generalized disease. In this study, CAD was identified as a significant risk factor based on the stepwise Cox PH model. Accordingly, patients with a known history of CAD before the diagnosis of NPC were assigned a score of 3 in our prediction model, reflecting the elevated risk associated with pre-existing atherosclerotic disease. This finding is also consistent with previously published literature.

On the other hand, the worldwide prevalence of obesity has increased in the past decades. Obesity, as defined by the World Health Organization (WHO), refers to an excessive accumulation of body fat that poses a risk to health. It is clinically diagnosed when an individual’s BMI is equal to or greater than 30 kg/m². Obesity is a well-established risk factor for numerous chronic conditions, including cardiovascular disease, type 2 diabetes, and certain cancers^38,39^. In addition to its association with metabolic dysregulation, obesity may also be linked to dietary habits and lifestyle factors. Unhealthy eating patterns and sedentary behaviors—such as prolonged sitting and lack of physical activity—can contribute to the development of conditions such as high blood pressure^40^ and insulin resistance^41^, thereby further increasing the risk of cardiovascular diseases. Nevertheless, the impact of obesity on carotid atherosclerosis disease is still unclear^42^. A systematic review found no significant association between overall obesity and CAS; however, the distribution of adipose tissue—particularly the ratio of visceral to subcutaneous fat—may play a role in the development and prevalence of carotid plaque^43^. Excess visceral fat is correlated with systemic inflammation and vascular inflammation^44,45^. Although a BMI of 30 or above is commonly used as the threshold for obesity according to the WHO, previous studies have shown that individuals of Asian descent tend to have higher body fat percentages at lower BMI levels compared to Western populations^46^. Accordingly, it has been proposed that lower BMI cutoffs be used to define obesity in Asian populations^47^. In the Taiwanese context, a BMI ≥30 often reflects not only general obesity but also significant visceral adiposity^48^, which is closely associated with metabolic and cardiovascular risks. Despite this ethnic-specific consideration, our study adhered to the WHO BMI classification. This may partly account for why our risk prediction model assigned a relatively higher risk score of 3 to individuals with a BMI of 30 or above, reflecting the substantial ICA stenosis risks associated with this level of obesity in our study population.

In addition, consistent with previous research^49,50^, smoking was identified as an independent risk factor for ICA stenosis in our cohort, further underscoring the need for comprehensive risk factor management in post-radiation NPC patients.

### The performance of our risk predication model

The model achieved an AUC of 0.623, indicating modest discriminatory ability in distinguishing between patients who did and did not develop moderate or greater ICA stenosis. Although this level of discrimination is not considered high, it remains clinically informative. Additionally, the predictiveness curve (Figure 4) showed that predicted risks were concentrated in the upper percentiles, indicating that the model is capable of identifying a high-risk subgroup for potential targeted interventions. More importantly, the IBS of risk prediction model was 0.084, reflecting excellent overall prediction accuracy throughout the study period. Given that the IBS incorporates both discrimination and calibration over time, a value below 0.1 suggests the model’s predicted probabilities align well with observed outcomes across the entire follow-up period. Together, these metrics suggest that while the model’s risk stratification may be moderate, its probabilistic forecasts are highly reliable, supporting its potential utility in guiding long-term surveillance strategies for high-risk patients

### Strength of this study

To the best of our knowledge, this is the first study to specifically investigate the risk factors associated with CAS specifically in NPC patients who have undergone radiotherapy. First, we aimed to develop a risk prediction model to identify high-risk individuals who may benefit from regular surveillance and potentially earlier intervention to prevent ischemic stroke. Second, our focus was to determine the optimal timing for initiating carotid ultrasound screening and surveillance. According to our findings, individuals in the high-risk group (with a risk score of 5 or above) may experience this occurrence as early as the fourth year after RT. Based on these results, we recommend that carotid Doppler US be initiated at the seventh-year post-radiation therapy for NPC patients who had underwent RT, while high-risk individuals should begin screening as early as the fourth year. This model enables precise and efficient risk stratification, guiding the implementation of individualized follow-up strategies, reducing unnecessary testing, and ultimately minimizing healthcare and time costs.

### Limitation of this study

There are several limitations in this study that need to be discussed. Firstly, this study is based on data from a single medical center so the sample size of our study is relatively small. Hence, future studies may require data collection from multiple institutions or nationwide sources to allow for broader analysis and validation.

Secondly, this study did not account for all potential risk factors related to CAS in the analysis. Important biochemical markers such as triglycerides, LDL, C-reactive protein, uric acid and homocysteine were not included for analysis. Although we included hyperlipidemia as a variable in the model, the level of disease control varies among patients. Utilizing biochemical markers such as triglycerides or LDL might provide a better assessment. Similarly, although chronic diseases such as hypertension and diabetes are included as variables in risk prediction models, there is considerable variability among patients in terms of disease duration and the degree of control over blood pressure and blood glucose. In prior studies investigating the prediction of CAS, patients with hypertension were stratified into eight groups according to antihypertensive medication use and systolic blood pressure levels^51,52^. The subgroup receiving anti-hypertension medication but with systolic blood pressure ≥160 mmHg exhibited the highest risk of moderate or more severe CAS. Therefore, incorporating objective measurements such as systolic blood pressure values or glycated hemoglobin (HbA1c) levels may provide a more accurate representation of patients’ actual clinical status. Furthermore, due to the relatively small number of participants included in the study, we were unable to investigate whether a dose-response relationship exists between smoking intensity or duration and the development of ICA stenosis. Future studies may consider integrating the dose-response relationship of smoking, as well as relevant biochemical markers, into risk prediction models to potentially improve predictive accuracy.

Thirdly, the outcomes of this study were determined based on carotid Doppler US, which may be subject to measurement variability and subjective interpretation. Nonetheless, all carotid US results in this study were assessed by experienced neurologists to minimize such errors. Additionally, the actual onset of ICA stenosis may have occurred prior to the time of examination, potentially causing the recorded timing of stenosis onset to be later than its true occurrence. In other words, the precise timing for carotid US screening to detect ICA stenosis in post-RT NPC patients may need to be earlier. Future studies with more rigorous experimental designs are warranted to confirm this.

Fourthly, despite the exclusion of cases with documented disease recurrence, loss to follow-up due to competing causes of death, such as unexpected mortality from NPC or other comorbidities, may have introduced bias and affected the validity of the study findings. Furthermore, the relatively small sample size limited our ability to adequately adjust for competing risks of death in the analysis, which could influence the robustness of the results.

Finally, since the study population consisted exclusively of Taiwanese individuals, the generalizability of the findings to other populations may be limited. Nonetheless, it is crucial to acknowledge that NPC is a notably high prevalence in Taiwan, posing a significant public health concern.

## CONCLUSION

In this study, NPC patients who underwent RT (mean age 48.5) had a 14.5% incidence of moderate to severe ICA stenosis over eight years—substantially higher than age-matched general population rates. A risk prediction model identified high-risk individuals, with those scoring ≥5 showing a significantly higher incidence, often beginning as early as four years post-radiotherapy.

The overall prediction accuracy of our risk predication model was excellent (IBS=0.084), and it provides two important insights. First, in patients with NPC who have undergone radiation therapy, the history of obesity (BMI ≥ 30), CAD, and smoking habits were found to be high-risk factors for developing carotid atherosclerosis sequelae in the future. These risk factors have a higher risk level than the chronic diseases we are more familiar with, such as hypertension, diabetes, and hyperlipidemia. This suggests that, after being diagnosed with NPC and beginning treatment, it is crucial not only to control hypertension, diabetes, and hyperlipidemia, but also to strongly advise patients to quit smoking and manage their weight, especially in male patients and those over 50 years old. Second, we recommend starting regular carotid Doppler US surveillance in the seventh year after radiation therapy. However, for high-risk individuals with a risk score above five, it is advisable to begin regular carotid doppler US from the fourth year. The surveillance recommendations proposed in this study warrant further validation. Moreover, other conditions such as autoimmune vasculitis, small vessel disease, and atrial fibrillation may also play a role in the pathogenesis of ischemic stroke. Consequently, further research is warranted to investigate whether early screening and timely intervention can effectively lower the incidence of ischemic stroke in NPC survivors.

## Data Availability

nil

## Research Funding

Not applicable

## Author Contributions

**Conception and Design:** Chuan-Yi Lin, MD; Chun-Nan Chen, MD, PhD; Po-Hsiu Kuo, PhD

**Collection and Assembly of Data:** Chuan-Yi Lin, MD; Chun-Nan Chen, MD, PhD

**Data Analysis and Interpretation:** Chuan-Yi Lin, MD; Szu-Yuan Wu, MD, MPH, PhD; Po-Hsiu Kuo, PhD

**Administrative Support:** Chun-Nan Chen, MD; Jenq-Yuh Ko, MD, PhD; Szu-Yuan Wu, MD, MPH, PhD

**Manuscript Writing:** Chuan-Yi Lin, MD; Chun-Nan Chen, MD; Po-Hsiu Kuo, PhD

**Final Approval of Manuscript:** All authors

